# Linking gene expression patterns and brain morphometry to trauma and symptom severity in patients with functional seizures

**DOI:** 10.1101/2021.12.01.21266852

**Authors:** Johannes Jungilligens, Stoyan Popkirov, David L. Perez, Ibai Diez

**Affiliations:** Department of Neurology, University Hospital Knappschaftskrankenhaus, Ruhr University Bochum, Bochum, Germany; Functional Neurological Disorder Unit, Division of Cognitive Behavioral Neurology, Department of Neurology, Massachusetts General Hospital, Harvard Medical School, Boston, MA, USA; Division of Neuropsychiatry, Department of Psychiatry, Massachusetts General Hospital, Harvard Medical School, Boston, MA, USA; Athinoula A. Martinos Center for Biomedical Imaging, Massachusetts General Hospital, Harvard Medical School, Boston, MA, USA; Gordon Center for Medical Imaging, Department of Radiology, Massachusetts General Hospital, Harvard Medical School, Boston, MA, USA

**Keywords:** functional neurological disorder, psychogenic non-epileptic seizures, trauma, symptom severity, gray matter, neuroimaging genetics

## Abstract

**Objective:** Adverse life experiences (ALEs) increase the susceptibility to functional (somatoform/dissociative) symptoms, likely through neurodevelopmental effects. This analysis aimed to illuminate potential genetic influences in neuroanatomical variation related to functional symptoms and ALEs in patients with functional seizures.

**Methods:** Questionnaires, structural brain MRIs and Allen Human Brain Atlas gene expression information were used to probe the intersection of functional symptom severity (Somatoform Dissociation Questionnaire, SDQ-20), ALE burden, and gray matter volumes in 20 patients with functional seizures.

**Results:** Functional symptom severity positively correlated with the extent of sexual trauma, emotional neglect, and threat to life experiences. In voxel-based morphometry analyses, increased SDQ-20 scores related to decreased bilateral insula, left orbitofrontal, right amygdala, and perigenual and posterior cingulate gray matter volumes. Left insula findings held adjusted for psychiatric comorbidities. Increased sexual trauma burden correlated with decreased right posterior insula and putamen volumes; increased emotional neglect related to decreased bilateral insula and right amygdala volumes. The sexual trauma–right insula/putamen and emotional neglect– right amygdala relationships held adjusting for individual differences in psychiatric comorbidities. When probing the intersection of symptom severity and sexual trauma volumetric findings, genes overrepresented in adrenergic, serotonergic, oxytocin, opioid, and GABA receptor signaling pathways were spatially correlated. This set of genes was over-expressed in cortical and amygdala development.

**Conclusion:** ALEs and functional symptom severity were associated with gray matter alterations in cingulo-insular and amygdala areas. Transcriptomic analysis of this anatomical variation revealed a potential involvement of several receptor signaling pathways.

## 1. Introduction

Adverse life experiences (ALEs) increase the risk for developing psychopathologies[1, 2]. Research understands this relationship to be driven by the effects of trauma on brain maturation and experience-dependent neuroplasticity, influenced by genetic/epigenetic, hormonal, neuroimmune, and social/environmental factors[3-6]. Across mood, anxiety and trauma-related disorders, ALEs are associated with an earlier onset, greater symptom severity and a more treatment refractory course[3, 7]. Additionally, discrete trauma subtypes have been linked to distinct brain alterations – providing additional specificity to these risk factor-brain organization relationships[8]. While less well-researched than other trauma-related disorders, functional neurological disorder (FND) is a neuropsychiatric condition where ALEs have been posited to be etiologically important[9-11]. Given the high prevalence and healthcare costs attributed to this population, FND is a model disorder through which to advance our neurobiological understanding of brain-trauma relationships across a range of psychopathologies[12, 13].

Functional seizures (FS), also known as dissociative or psychogenic nonepileptic seizures, are one of the most common and disabling FND subtypes[14]. Patients with FS experience paroxysmal events resembling epileptic seizures, yet this condition is not explained by electrical neural discharges and has distinct clinical features[15]. While not universally present, elevated ALE rates have been reported in patients with FS – with an odds ratio of 3.1 compared to controls[9]. Selkirk and colleagues identified that patients with FS reporting antecedent sexual trauma exhibited an earlier illness onset, more severe convulsions, and worse overall psychiatric health[16]. Positive associations between the magnitude of previously experienced ALEs and symptom severity have been independently replicated in several other mixed-FND cohorts[17-19]. This “dose-dependent” relationship between ALEs and symptom severity underscores the etiological and likely mechanistic relevance of traumatic experiences in the pathophysiology of FS. The magnitude of ALEs in FS cohorts also correlated with biased attentional/threat processing, dissociation, maladaptive coping, and altered neuroendocrine profiles – further underscoring the relevance of ALEs in this population[20-24]. In a mixed-FND cohort, sexual abuse was linked to poor treatment outcome[25] – highlighting additional prognostic implications.

Several studies used structural brain magnetic resonance imaging (MRI) to investigate gray matter profiles in patients with FS. While the direction of the effect has been inconsistent across studies[13, 26], cingulo-insular, medial orbitofrontal, lateral prefrontal, and amygdalar volumetric alterations have been identified in FS cohorts compared to healthy controls (HCs)[27-31]. Two studies showed that symptom severity negatively correlated with medial orbitofrontal and inferior frontal volumes in FS cohorts[28, 29]. Similarly, in mixed-FND including FS, cingulo-insular atrophy correlated with individual differences in patient-reported symptom severity[19, 32]. Importantly, most FS cohort studies did not account for the magnitude and range of previously experienced ALEs, and did not evaluate relationships between symptom severity, ALEs, and gray matter profiles. Given that there may be both disease-related and non-specific neuroplastic changes related to the magnitude of ALEs – including biologically relevant consequences of discrete trauma subtypes[8, 33, 34] – these are important gaps that if clarified would help advance our pathophysiological understanding of FS[35], and enrich our knowledge of transdiagnostic risk factors.

In addition to quantitative structural MRI analyses, investigating the intersection of neuroimaging-derived phenotypes (e.g., gray matter statistical maps) and gene expression profiles offers a cutting-edge complementary approach to characterize neurobiological disease mechanisms[26, 36]. Spatial correlations between neuroimaging findings and brain-wide gene expression atlases (e.g., Allen Human Brain Atlas, AHBA) allow inferences about genes over-expressed in implicated brain regions without the need for individual genotyping[37]. This offers a valuable platform to understand not only the genetic influences on brain structure, but also the cellular-molecular signaling pathways with potential disease relevance. Positron emission tomography (PET)-based work found that AHBA-based gene-expression patterns are representative of actual receptor density, highlighting the validity of neuroimaging-genetic approaches[38]. Such research has provided valuable mechanistic insights regarding neuroplasticity across a range of neuropsychiatric and healthy populations[39, 40]. In FND, research into potential genetic factors is in its infancy. Genetic studies performed to date provide early support for a mechanistic role for neurotransmitter pathways (e.g., oxytocin, serotonin) and brain development more broadly[41-43].

Here, we first aimed to examine the relationship between specific ALE subtypes (e.g., sexual trauma, emotional neglect etc.), patient-reported symptom severity, and illness duration in patients with FS. Thereafter we examined relationships between gray matter volumes and indices of functional symptom severity, illness duration, and magnitude of previously experienced ALE subtypes. Thirdly, we tested for spatial similarities between imaging-derived phenotypes and AHBA gene expression profiles – with an interest in identifying genetic pathways dually implicated in the association of volumetric gray matter variations with symptom severity and trauma burden in patients with FS. This approach is consistent with the recently published FND neuroimaging research agenda proposed by international experts[44]. We hypothesized that the magnitude of ALEs with a direct physical component (i.e., sexual abuse, physical abuse) would most strongly correlate with symptom severity and illness duration[9, 16, 41, 45]. We also hypothesized that reduced gray matter volumes within salience and limbic network brain areas would relate to symptom severity and trauma burden. Lastly, when investigating relationships between imaging-derived phenotypes and gene expression profiles, we hypothesized that genes implicated in stress-related signaling pathways and brain development would be identified. As we specifically aimed to investigate potential transdiagnostic developmental pathways for the impact of ALEs on the development of brain-mind-body disorders using FS as a model condition, we do not include a healthy control group.

## 2. Patients and Methods

### 2.1. Participants

Twenty adults with FS (15 females, mean age 34.9±11.2 years, mean illness duration 5.2±5.3 years) were enrolled. Following established criteria, FS diagnoses were documented on video-EEG (n=13) or confirmed by expert clinical consensus (n=7)[46]. Exclusion criteria included major neurological comorbidity (e.g., epileptic seizures), abnormal brain MRI, epileptiform discharges on EEG, and/or inconclusive video-EEG evaluation. See **Supplementary Methods** for details on participant enrollment. A structured psychiatric interview (Mini-DIPS[47]) was used to screen all participants for lifetime psychiatric disorders. Fourteen of 20 patients with FS fulfilled criteria for lifetime major depressive disorder (MDD), 9/20 for panic disorder (PD), and 16/20 for post-traumatic stress disorder (PTSD). Four patients were on anti-depressants. Detailed demographic and clinical information is provided in **Supplementary Table 1**, and as previously reported for a diffusion tensor imaging analysis performed in this same cohort[48]. All participants provided written informed consent and this study was approved by the ethics committee of the medical faculty, Ruhr University Bochum (Reg.-Nr. 17-6019).

### 2.2. Self-Report Questionnaires

Functional symptom severity was quantified using the Somatoform Dissociation Questionnaire-20, a 20-item self-report measure with each item scored on a 5-point Likert scale (SDQ-20[49]; German version[50]).

The presence and severity of previously experienced ALEs across the lifespan was recorded using the Traumatic Experiences Checklist (TEC[51]; German version[52]). The TEC is a self-report measure of lifetime adverse and/or potentially traumatic experiences indexed across six subscales (emotional neglect, emotional abuse, physical abuse, threat to life, sexual harassment, and sexual abuse). Here, “sexual harassment” and “sexual abuse” sub-scores were nearly identical in our cohort (partial correlation r=.928, p<0.001; controlled for age, sex) – thus a combined “sexual trauma” score was used throughout.

Relationships between SDQ-20, illness duration, and TEC subscale scores were established using partial correlation analyses, controlling for age and sex. A p<0.05 was considered significant.

### 2.3. Structural Neuroimaging

#### Voxel-Based Morphometry Preprocessing

See **Supplementary Methods** for structural T1 acquisition parameters. FSL’s Voxel Based Morphometry (VBM[53, 54]) was used to characterize gray matter volumes. First, a study-specific gray matter template was created by brain-extracting and gray matter-segmenting all structural images before registering them to the MNI-152 standard space using non-linear FNIRT registration[55]. The resulting images were averaged and flipped along the x-axis to achieve left-right symmetry. Thereafter, all gray matter images were registered to this template (using FNIRT) and corrected for local expansion or contraction (due to the non-linear component of the spatial transformation). Smoothing with an isotropic Gaussian kernel with a sigma of 2mm was applied. To correct for nonspecific brain size influences, total intracranial volume was measured using FreeSurfer’s recon-all processing stream[56].

#### Individual Differences in Gray Matter Volumes

A single-class GLM was used to investigate relationships between individual differences in gray matter volumes and illness severity (SDQ-20 score, illness duration) or TEC subscores in patients with FS. All analyses controlled for age, sex, and total intracranial volume. Whole-brain, cluster-wise correction for multiple comparisons was applied using AFNIs Monte Carlo simulation with a cluster-forming threshold p<0.05 and 10,000 iterations. For statistically significant findings, post-hoc analyses controlled for comorbid psychopathology (lifetime MDD, PD, and/or PTSD for illness severity findings or lifetime MDD and/or PD for trauma-associated findings).

### 2.4. Exploratory Gray Matter – Gene-Expression Analyses

#### Imaging-Derived Gene Expression Profiles

To explore relationships between gray matter and gene expression profiles, we used a data-driven approach utilizing spatial similarity metrics between gray matter statistical maps (furthermore referred to as “imaging derived phenotypes”) and regional gene expression patterns (for methodological overview, see[36]). Individual imaging-derived phenotypes from above-described within-group VBM analyses of relationships between gray matter volume and SDQ-20 or TEC subtypes were used to investigate spatial similarities with AHBA gene expression data[37]. The AHBA provides high-resolution, whole-brain genome-wide expression values obtained from six human donors across 20,736 genes in 3,702 samples distributed throughout the brain. After averaging gene expression of multiple probes, we projected all brain sample expression values to 84 (68 cortical, 16 subcortical) Desikan atlas brain regions[57]. For each donor, the median gene expression of all samples within each region was computed. To obtain a unique brain map for each gene, the average expression between different donors was computed. Given a specific interest in genes associated with neurobiological structure and function, we focused on 2,382 brain-related genes that overlapped between the genes mapped in the AHBA and those having an at least fourfold elevated expression in the brain compared to other organs[58].

For each imaging-derived phenotype, the t-statistical value of each brain area was extracted and the Spearman rank correlation between regional t-statistic vectors and all brain-related genes was computed. Specifically, the statistical maps of the relationship between gray matter volume and 1) SDQ-20, 2) TEC-emotional neglect, and 3) TEC-sexual trauma were separately correlated with spatial AHBA gene expression maps. Genes with a spatial similarity >2 standard deviations from the mean were extracted. Genes in the lower tail of the distribution were analyzed to identify genes for which higher expressions correlated with lower volume. Furthermore, to remove variability in these respective gene sets, we used K-means clustering to identify gene groups with similar expression values. Silhouette algorithms were used to identify the optimal number of clusters.

#### Gene-Set Enrichment Analyses

Identified gene sets were further analyzed using gene-set enrichment[40]. Enrichment analyses search for functional, structural, and/or process annotations that are statistically overrepresented in a specific set of genes to aid interpretation of the implications. Here, we used PANTHER pathways version 16[59], accessed through gene-ontology.org. Fisher’s exact test and FDR correction p<0.05 was used.

Additionally, to explore the neurodevelopmental implications of the identified gene sets, we used the Specific Expression Analysis tool (SEA[60]; http://genetics.wustl.edu/jdlab/csea-tool-2). SEA explores the neurodevelopmental effects of genes within a given set by analyzing the extent to which they are involved in the formation and differentiation of certain brain structures (i.e., amygdala, cerebellum, cortex, hippocampus, striatum, thalamus) across different developmental stages (from early fetal to young adulthood). The specificity of the genetic annotations is indicated by a specificity index (pSI), with a smaller pSI representing higher structure-specific levels of a small number of genes, and a higher pSI relating to a larger number of relatively enriched genes. Relationships are tested using Fisher’s exact test with Benjamini-Hochberg correction for multiple comparisons.

## 3. Results

### 3.1. Relationships Between Illness Severity and Trauma Burden

In patients with FS, SDQ-20 scores positively correlated with the magnitude of previously experienced emotional neglect (r=0.48, p=0.045), sexual trauma (r=0.55, p=0.018), and threat to life (r=0.76, p=0.001), see **Table 1**. Illness duration did not significantly correlate with any trauma subtype. See **Supplementary Table 2** for additional details regarding questionnaire scores.

**Table 1.** Partial correlations (age, sex adjusted) between illness severity measures and trauma subtype scores in patients with functional seizures.

|  | TEC<br>Emotional<br>neglect | TEC<br>Emotional<br>abuse | TEC<br>Physical<br>abuse | TEC<br>Threat to<br>life | TEC<br>Sexual<br>trauma |
| --- | --- | --- | --- | --- | --- |
| SDQ-20 score | **0.48**<br>(**$p=0.045$**) | 0.18<br>($p=0.48$) | -0.12<br>($p=0.65$) | **0.76**<br>(**$p<0.001$**) | **0.55**<br>(**$p=0.018$**) |
| Illness duration | -0.070<br>($p=0.78$) | 0.054<br>($p=0.84$) | 0.18<br>($p=0.49$) | -0.41<br>($p=0.099$) | 0.38<br>($p=0.12$) |

**Association between trauma burden subtypes and functional symptom severity in patients with functional seizures (FS).** All associations were established using partial correlations controlling for age and sex. Strength of correlation (partial correlation coefficients) is indicated for each analysis. Trauma burden was measured using the Traumatic Experiences Checklist (TEC) and its respective subscales; functional symptom severity was assessed using the Somatoform Dissociation Questionnaire (SDQ-20). Significant correlations ( $p < 0.05$ , marked in bold) were found for emotional neglect, threat to life, and sexual trauma. Abbreviations: SDQ-20=Somatoform Dissociation Questionnaire-20, TEC=Traumatic Experiences Checklist.

### 3.2. Individual Differences Between Gray Matter Volumes and Illness Severity

In patients with FS, elevated SDQ-20 scores were associated with reduced bilateral insula, perigenual anterior cingulate cortex (ACC), posterior cingulate cortices, right amygdala, dorsal precentral/motor, left orbitofrontal cortex, ventral precentral gyrus, and brainstem volumes (**Figure 1A**). When controlling for lifetime MDD, PD, PTSD, the left insula/ventral precentral gyrus cluster remained significant.

**Figure 1.**
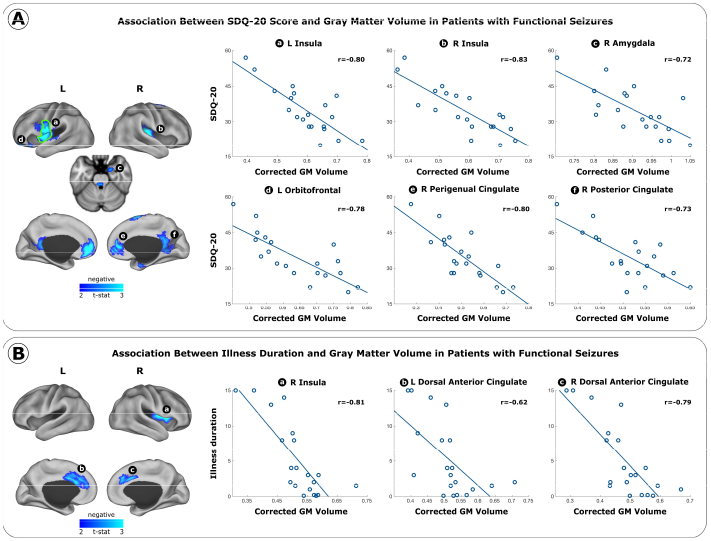
Gray matter volume associations with illness severity measures in patients with functional seizures (FS). A general linear model was used to study associations controlling for age, sex, total intracranial volume. Significant clusters surviving to multiple comparisons are colored by the direction of effect (blue: negative association). Scatterplots display relationships between corrected gray matter volumes and illness severity. Strength of correlation (partial correlation coefficients) is indicated for each cluster. Clusters remaining significant following multiple comparisons correction when additionally adjusting for lifetime MDD, PD, and PTSD are highlighted using green borders. **Panel A** displays the relationship between SDQ-20 scores and gray matter volumes. **Panel B** displays the relationship between illness duration and gray matter volumes. Abbreviations: L=Left, R=Right, GM=Gray Matter, SDQ-20=Somatoform Dissociation Questionnaire-20.

Regarding illness duration, reduced bilateral dorsal ACC and right insula volumes were associated with greater illness duration in patients with FS (**Figure 1B**). When controlling for lifetime MDD, PD, PTSD, no clusters remained significant.

### 3.3. Individual Differences Between Gray Matter Volumes and Trauma Burden

In patients with FS, individuals reporting greater emotional neglect showed relative reductions in bilateral insula and right amygdala volumes as well as positive associations with left lateral occipital volumes (**Figure 2A**). The right amygdala and left occipital clusters remained significant when controlling for lifetime MDD and PD. In a separate analysis, patients with FS with higher sexual trauma scores showed relative reductions in right insula and putamen volumes as well as inceases in right occipital/posterior temporal areas (**Figure 2B**). The right insula and putamen clusters remained significant when controlling for lifetime MDD and PD. There were no statistically significant correlations between gray matter volumes and threat to life trauma scores in patients with FS.

**Figure 2.**
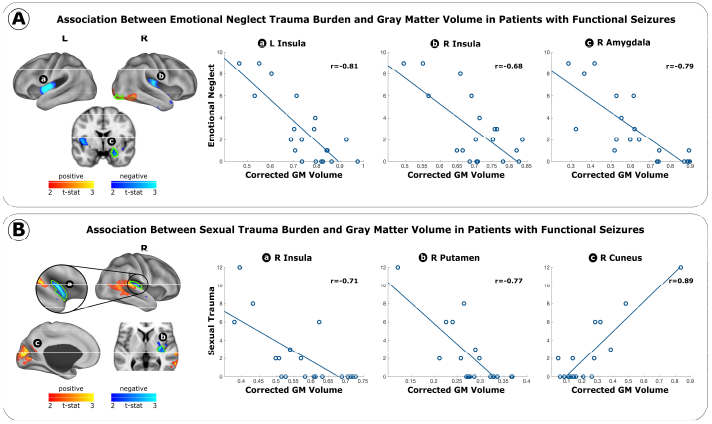
Gray matter volume associations with trauma burden subtypes in patients with functional seizures (FS). A general linear model was used to study associations controlling for age, sex, and total intracranial volume. Significant clusters surviving multiple comparisons are colored by the direction of effect (blue: negative correlation). Scatterplots display relationships between corrected gray matter volumes and trauma burden. Strength of correlation (partial correlation coefficient r) is indicated for each cluster. Clusters remaining significant when additionally correcting for lifetime MDD and PD are highlighted using green borders. **Panel A** displays the relationship between corrected gray matter volumes and emotional neglect scores in patients with FS. **Panel B** displays the relationship between corrected gray matter volume with sexual trauma scores in patients with FS. Abbreviations: L=Left, R=Right, GM=Gray Matter.

### 3.4. Gray Matter – Gene-Expression Findings

Exploring gray matter – gene-expression relationships, the spatial similarity of the imaging-derived phenotypes (i.e. SDQ-20, emotional neglect, sexual trauma) and regional gene-expression profiles from the AHBA (**Figure 3A**) revealed two clusters per phenotype (**Figure 3B left**, see **Supplementary Table 3** for the complete lists of genes). In gene-set enrichment analyses for PANTHER pathways, only one of two clusters per variable reached statistical significance (**Figure 3B right**).

**Figure 3.**
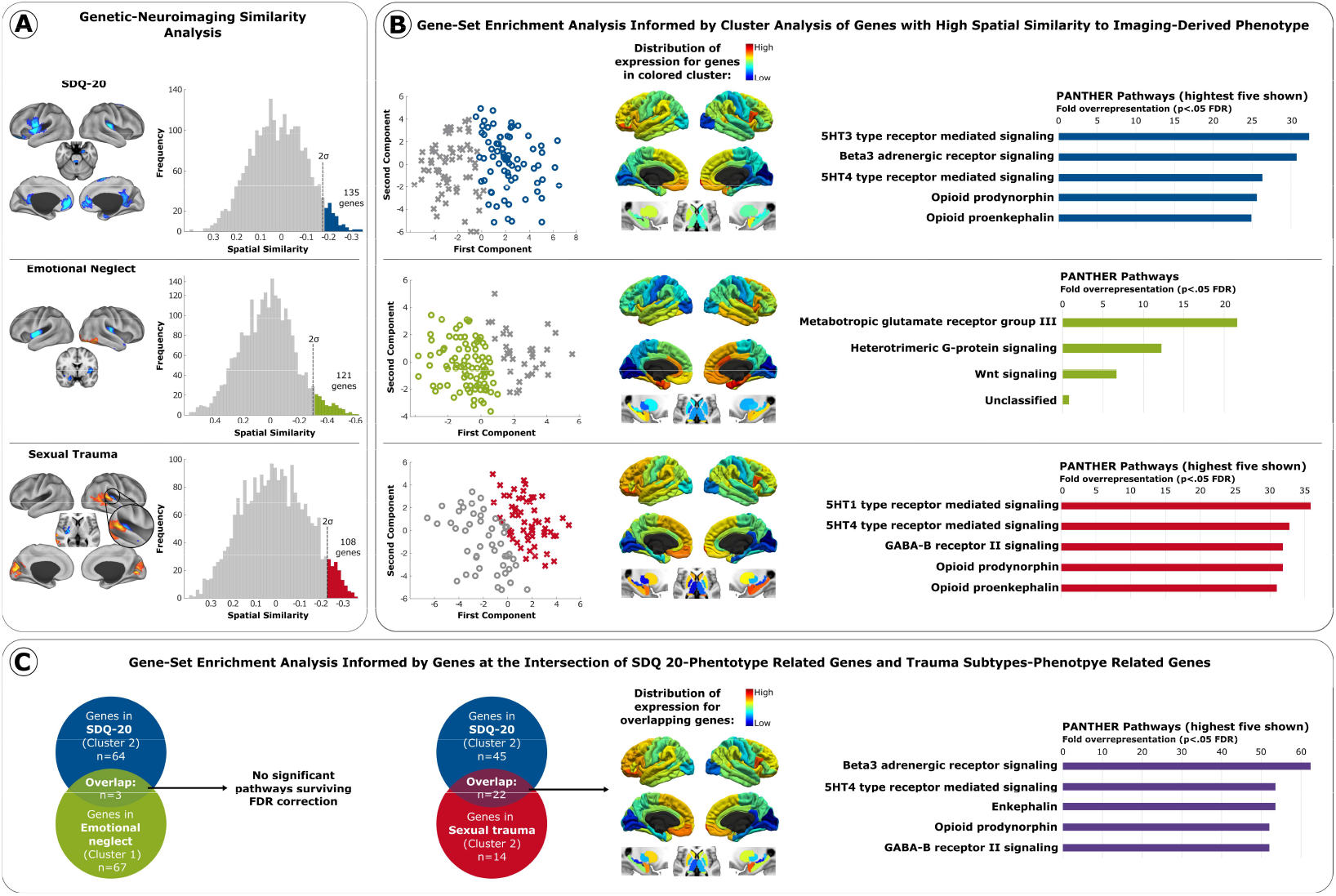
Spatial correlations between imaging-derived phenotypes and Allen Human Brain Atlas (AHBA) gene-expression profiles. Panel. **A** displays the spatial similarity analysis: imaging-derived phenotypes (t-statistic maps) for the associations between gray matter volume and 1) SDQ-20 (upper panel A), 2) emotional neglect (middle panel A), and 3) sexual trauma (lower panel A) were individually correlated with spatial expression maps from the AHBA. Genes with a spatial similarity >2 standard deviations (SD) from the mean in the lower tail of the distribution were extracted. **Panel B** displays the gene-set enrichment: cluster analysis revealed two clusters per spatial similarity distribution (left side of panel B). The distribution of gene expression for the genes from each colored cluster was mapped to enable a cross-reference to the original imaging-derived phenotypes (middle of panel B). The list of genes for each cluster were entered into gene-set enrichment analyses for PANTHER pathways (right side of panel B). Only one of two clusters per variable (i.e., SDQ-20, emotional neglect, sexual trauma) reached statistical significance in pathway analyses. Top five pathways for each condition are shown, see **Supplementary Table 4** for full list. **Panel C** displays the overlap analysis: Informed by the above-mentioned gene-sets, lists of genes which overlapped between 1) SDQ-20- and emotional neglect-derived maps, and 2) SDQ-20- and sexual trauma-derived maps were entered into gene-set enrichment analysis to investigate common pathways. Top five pathways (with highest fold overrepresentation) for each condition are shown, see **Supplementary Table 4** for full list. Additional visualization for gene-expression for positive tail of sexual trauma are provided in **Supplementary Figure 1**.

For brain regions with spatial relationships with SDQ-related gray matter volumes, pathways with the highest fold overrepresentation (FDR-corrected significant) included serotonergic, adrenergic, and opioid receptor signaling (**Figure 3B, upper panel**; full list in **Supplementary Table 4**). Regarding the emotional neglect-related gray matter phenotype, we found an FDR corrected overepresentation of genes related to pathways for glutamate receptor, G-protein receptor, and Wnt signaling pathways (**Figure 3B, middle panel**). Lastly, regarding the sexual trauma-related gray matter phenotype, amongst the highest FDR corrected overrepresentation of genes were pathways related to serotonin, GABA, and opioid receptors (**Figure 3B, lower panel**; full list in **Supplementary Table 4**).

Next, we checked for genes that overlapped between the SDQ-related imaging-derived phenotype and trauma subtype-related imaging-derived phenotypes (individually for emotional neglect and sexual trauma, **Figure 3C**). For the overlap between gene sets in SDQ-related imaging-derived phenotypes and emotional neglect-related imaging-derived phenotypes (three genes), no pathways were found (**Figure 3C left**). On the other hand, 22 genes overlapped between the gene set from SDQ-related imaging-derived phenotypes and the gene set from sexual trauma-related imaging-derived phenotypes (see **Supplementary Table 3** for full list). These 22 genes showed the highest statistical overrepresention in pathways related to adrenergic, serotonergic, enkephalin, opioid, and GABA receptor signaling (**Figure 3C right**, see **Supplementary Table 4** for full list).

Using the CSEA Specific Expression Analysis to explore potential neurodevelopmental implications, the 22 overlapping genes (SDQ-20 x sexual trauma) were identified to be related to cortical maturation (from neonatal to young adulthood) and amygdala development in adolescence and young adulthood (**Figure 4**). Furthermore, they were enriched in prenatal thalamic development.

**Figure 4.**
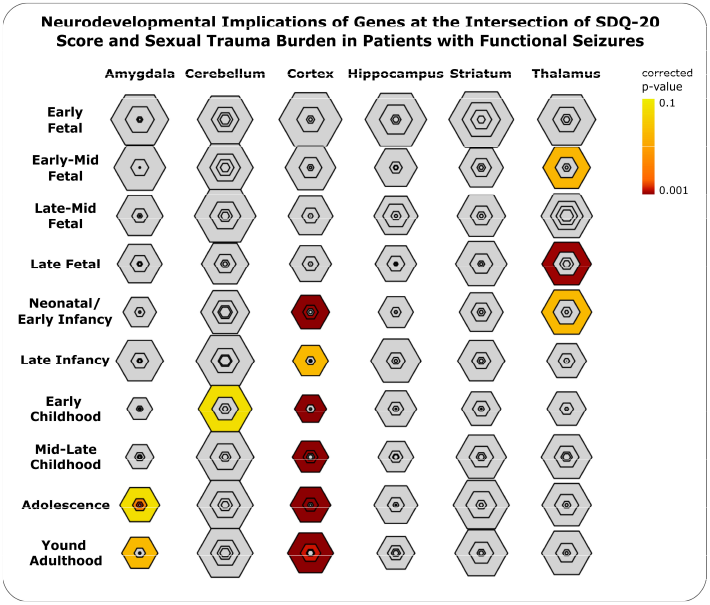
Neurodevelopmental implications of enriched gene sets. The sets of genes overlapping between SDQ-20- and sexual trauma-derived maps were entered into a gene-set enrichment analysis with neurodevelopmental annotations (SEA tool, http://genetics.wustl.edu/jdlab/csea-tool-2) to investigate implications of these genes across brain regions and developmental stages. Smaller/more central hexagons imply higher structure-specific gene annotations; larger/less central hexagons imply larger sets of related genes. Darker colors relate to higher Benjamini-Hochberg corrected significance for the association between the gene-set and a specific region in a specific developmental stage. i.e., for a given region with three hexagons (e.g., amygdala in adolescence). The enrichment analysis is moderately significant for a larger set of related genes (outer, bright-yellow hexagon), and additionally highly significant for higher structure-specific set of genes annotations (inner, orange-red hexagons).

## Discussion

This study identified several key findings: i) patient-reported functional symptom severity (SDQ-20 scores) positively correlated with the magnitude of specific ALE subtypes (sexual trauma, emotional neglect, threat to life). ii) Increased SDQ-20 scores related to decreased bilateral insula, left orbitofrontal, right amygdala, and perigenual and posterior cingulate gray matter volumes; individual differences in SDQ-20 scores correlated with reduced left insula/ventral precentral gyrus volumes held adjusting for lifetime MDD, PD and PTSD. iii) increased sexual trauma burden correlated with decreased right posterior insula and putamen volumes; increased emotional neglect related to decreased bilateral insula and right amygdala volumes. The sexual trauma– right insula/putamen and emotional neglect–right amygdala relationships held adjusting for individual differences in MDD and PD. iv) At the intersection of the SDQ-20 and the sexual trauma imaging-derived phenotypes, there was significant spatial correlation with genes overrepresented in adrenergic, serotonergic, oxytocin, opioid, and GABA receptor signaling pathways. v) The same set of genes related to cortical (from neonatal to young adulthood) and amygdala (from adolescence to young adulthood) development.

Increased illness severity (measured by SDQ-20 and illness duration) related to reduced cingulo-insular and orbitofrontal volumes. The correlation between left anterior insula volumes and SDQ-20 scores is particularly noteworthy given that this finding held adjusting for comorbid psychopathology. Consistent with this finding, relationships between increased functional symptom severity and decreased left anterior insula gray matter have previously been reported in a mixed-FND cohort[19, 61]. Relatedly, increased anterior insula – sensorimotor network connectivity correlated with seizure frequency in an FS cohort[62]. While not withstanding correction for comorbid psychopathology, our finding of orbitofrontal volume reductions related to higher SDQ-20 scores is also congruent with prior studies in FS cohorts[28, 29]. Additionally, across SDQ-20 score and illness duration analyses, relationships with reduced salience network (SN) and default mode network (DMN) brain areas were found, including the right amygdala and the dorsal, perigenual and posterior cingulate. These findings did not hold accounting for psychiatric comorbidities, suggesting that these brain areas may have shared pathophysiological implications across FS and other mood/anxiety/trauma-related disorders[63]. The SN is implicated in affective experiences and attention[64] and has been identified as highly relevant for FND pathophysiology[35, 65]. Specifically, the insula (involved in interoception and subserving emotion and self-awareness[66, 67]) and the ACC (a hub implicated in cognitive control, affect, and nociception[63]) has repeatedly been connected to the neuropathophysiology of FND across structural and functional imaging studies[32, 68-72]. Alterations in the DMN – involved in self-referential processing and information compression across modalities to enable conceptualization and contextualization of perceptions and experiences[73-75] – have also been linked to FND[13, 76, 77], MDD[78, 79] and PTSD[74, 80].

Given that functional symptom severity related to SN and DMN structural alterations, it is notable that sexual trauma and emotional neglect burden also correlated with reduced insula volumes in patients with FS; for sexual trauma, this finding held correcting for comorbid psychopathology. This suggests that discrete ALE subtypes may promote neuroplastic changes specifically contributing to a vulnerability for the development of FS later in life. This understanding fits well with the positive correlations between SDQ-20 and discrete trauma subtype scores found here, findings that replicate and extend the previously literature noting dose-dependent trauma – symptom severity relationships in FS and FND populations[17-19]. The overlap of trauma and symptom severity associated structural imaging findings has previously been reported in mixed-FND[19], but not in FS cohorts, and offers a bridge between etiological factors and disease mechanisms. From a broader perspective, our findings also advance the understanding of the range of aberrant neuroplastic change that can occur in the setting of adversity across clinical and healthy populations[8, 41, 81-83]. However, replication and longitudinal studies are needed to comprehensively understand how distinct ALE subtypes and features (e.g., the physicality of the trauma) are related to FND symptoms. Across a diverse range of psychopathologies, childhood sexual abuse is most strongly associated with FND[45]; however, a subset of patients with FND do not report prominent ALEs – underscoring the need to continue to investigate the full range of biopsychosocial-informed risk factors for FND and related disorders. Embedding results from FND cohorts into the wider field of research on trauma-related disorders is likely to offer the opportunity to fully contextualize the contributions and interplay of predisposing vulnerabilities, developmental periods, timing, type, and extent of ALEs, as well as related social-environmental factors[7, 84].

Having identified overlapping brain – SDQ-20 and brain – trauma relationships in our cohort, we subsequently tested for shared genetic influences and cellular-molecular signaling pathways of these imaging-derived phenotypes. We observed that symptom severity and sexual trauma-derived phenotypes mapped onto pathways implicated in serotonin, oxytocin, adrenalin, and opioid receptor signaling. Interestingly, two previous genotyping studies in motor-FND found i) that serotonin receptor gene polymorphism was related to symptom severity, childhood abuse burden, and functional amygdala – middle frontal gyrus connectivity, and ii) an increased methylation of the oxytocin receptor gene[42, 43]. Overall, genetic variations in serotonin and oxytocin receptor systems have been related to amygdala reactivity, acute stress responses, and affective vulnerabilities to everyday events[4, 6, 85-89]. Type and severity of stressors has been shown to differentially influence region-specific serotonin receptor gene expression (e.g., chronic psychosocial stress reduced expression in cingulo-insular and hippocampal regions[90]), further corroborating the importance of ALE subtyping[91]. Combined with the existing literature, our study adds to the growing evidence of a role for serotonin and oxytocin receptor signaling pathways at the intersection of ALEs, symptom severity, and FND neurobiology. However, it is unlikely that these gene variations are FND-specific, but rather represent genetic susceptibilities that, combined with life stressors and other risk factors, may lead to individual FND phenotypes. Large-scale efforts involving individual genetic and epigenetic data, neuroimaging, ALE subtyping, and symptom severity measures are needed to further disentangle these factors.

Lastly, we sought to contextualize the neurodevelopmental implications of genes that overlapped between SDQ-20 and sexual trauma-related imaging-derived phenotypes. Genes were involved in development and maturation of i) the amygdala in adolescence and young adulthood, ii) cortical areas from neonatal age to young adulthood, and iii) thalamus in prenatal stages. Neurodevelopment and neuroplasticity related genes have previously been identified in mixed-FND: spatial expression profiles of the brain-derived neurotrophic factor gene correlated with physical abuse burden-dependent functional connectivity patterns[41]. Together with our results, this underscores the potential role of ALEs and their differential effects in developmental periods, which likely contributes to the heterogeneity found across FND studies (e.g., studies on amygdala volume in FND including FS have shown both increases and decreases[31, 92]). Studies have previously shown that ALEs in childhood are much stronger associated with FND than ALEs in adulthood, but a more fine-grained analysis of the timing of ALEs and the relationship to critical periods for neuroplasticity that may predispose to altered brain development is warranted. Lastly, the potential role of prenatal influences in the context of inter-generationally transmitted stress should be explored.

Within a transdiagnostic framework, ALEs play a pivotal etiological role in the development of psychopathology, including in brain-mind-body disorders such as FND. The impact of ALEs on developmental trajectories potentially leading to concurrent psychiatric disease is well-reflected in this study. Most FS patients in our sample report the presence of ALEs, and a majority (16/20) fulfilled criteria for lifetime diagnosis of PTSD. We specifically understand that associations between illness severity and gray matter volumes are influenced by a range of factors including ALEs – so the fact that VBM findings do partially not hold when correcting of comorbid psychopathology including PTSD is not surprising. This might in part be due to reduced variance and thus lower statistical power to detect significant associations through the inclusion of additional covariates, but also reflects the multifactorial contributions to the measured phenotype.

This study has several limitations, including modest sample size, which raises concerns about replicability in larger samples. However, our results are congruent with previous findings in FS and mixed-FND cohorts. Furthermore, comorbid psychopathology (especially MDD, PD, PTSD) was assessed as lifetime diagnoses based on a structured psychiatric interview, but not using dimensional questionnaires. Patients did not complete a weekly seizure frequency diary – which could have been used as a more specific measure of FS symptom severity[93]. As the study aimed to elucidate fundamental relationships between lifetime trauma burden, brain morphology, and symptom severity using FS as a model disorder, the inclusion of a psychiatric control group would have enhanced information on the disorder-specificity of our findings[29, 41]. This is the first study to explore gray matter–gene expression relationships in FND, and thus requires replication in independent samples – including the future inclusion of trauma and psychiatric controls. Future efforts can also aim to investigate the relevance of critical development periods, and their role in predisposing to the development of FS and related conditions. Lastly, there is a need to further characterize etiological factors and neural mechanisms in patients with FS and other FND subtypes that do not endorse a history of ALEs.

In conclusion, individual differences in symptom severity and trauma burden were associated with gray matter alterations in cingulo-insular and amygdala areas in patients with FS. Exploratory neuroimaging-genetics analyses underscored the potential role of genes involved in stress-related signaling pathways (serotonin, oxytocin) and neurodevelopment in the pathophysiology of FND. These study underscores the clinical and neurobiological importance of investigating a trauma subtype of FS and FND more broadly – while also maintaining a focus on the study of individual differences.

## Data Availability

Anonymized data and analyses scripts will be shared with qualified researchers contingent on approval by the local ethics committee.

## Ethical Standards

The authors assert that all procedures contributing to this work comply with the ethical standards of the relevant national and institutional committees on human experimentation and with the Helsinki Declaration of 1975, as revised in 2008. This study was approved by the ethics committee of the medical faculty, Ruhr University Bochum (Reg.-Nr. 17-6019). All participants provided written informed consent

