## Supplementary Methods for "Linking gene expression patterns and brain morphometry to trauma and symptom severity in patients with functional seizures"

### PARTICIPANT ENROLLMENT

Twenty adults with functional seizures (15 females) were prospectively recruited over 16 months from the inpatient and outpatient clinic of a tertiary epilepsy center at a university hospital in Germany. Diagnoses were made by a specialist team. Based on established criteria from the International League Against Epilepsy (LaFrance et al., 2013), diagnoses were documented on video-EEG (n=13) or confirmed by expert clinical consensus (n=7). Exclusion criteria for patients included neurological comorbidity (e.g., epileptic seizures), abnormal brain MRI, epileptiform discharges on EEG, and/or inconclusive video-EEG evaluation.

Screening for psychiatric diagnoses was performed using a structured diagnostic interview (Mini-DIPS, Margraf et al., 2017).

#### Brain MRI acquisition

MRI data was acquired on a Siemens Magnetom Prisma 3T Scanner using a 64-channel phased-array head coil. Head motion was restricted using either sound-isolating headphones or foam pads. High resolution 3D T1-weighted images were acquired using a magnetization prepared rapid gradient-echo (MPRAGE) sequence with the following parameters: 0.85 mm isotropic voxels, 208 sagittal slices, acquisition matrix size=288x288 mm, repetition time (TR)=1800 ms, echo time (TE)=2.39 ms, field of view (FOV)=288 mm, flip angle (FA)=8°, slice thickness=0.85mm.
