## Supplementary Table 1 for "Linking gene expression patterns and brain morphometry to trauma and symptom severity in patients with functional seizures"

**Supplementary Table 1. Characteristics of Study Participants with Functional Seizures.**

| Age group (years) | Sex | Illness Duration | Psychiatric Diagnoses | CNS-Active Medications |
|---|---|---|---|---|
| 41-45 | F | 15 years | PD, AG, BP II, MD, PTSD, INS, ICD, SI, SHB | Valproate, Quetiapine |
| 41-45 | F | 13 years | PD, AG, SAD, HME, MD, ICD, INS, PSY, SI | Pregabalin, Quetiapine, Risperidone, Venlafaxine |
| 26-30 | F | 8 years | GAD, INS, SI | - |
| 21-25 | F | less than a year | MD | - |
| 41-45 | F | 2 years | INS | - |
| 26-30 | F | 9 years | PTSD, MD | - |
| 46-50 | M | less than a year | PD, SAD, MD, PDD | Quetiapine, Zopiclone |
| 26-30 | F | 14 years | MD, PTSD, INS | - |
| 46-50 | M | 3 years | PDD, AUD, INS | Trimipramine, Pregabalin |
| 21-25 | F | 2 years | GAD, MD, PTSD, INS, PSY, SHB | - |
| 21-25 | F | 1.5 years | AG, SP, GAD, MD, INS, SHB | Rotigotine |

| | | | | |
|---|---|---|---|---|
| 41-45 | M | 3 years | PD, SP, GAD, MD | Paroxetine |
| 36-40 | F | 1.5 years | PD, AG, SP, PDD, PTSD, ED, INS | Clonazepam |
| 26-30 | F | 4 years | GAD, INS | - |
| 21-25 | M | less than a year | - | - |
| 51-55 | M | 4 years | GAD, MD, PDD, ICD, SSD, AUD, INS, SI | - |
| 56-60 | F | 8 years | PD, GAD, MD, PDD, PTSD, INS | Trazodone, Amitriptyline |
| 21-25 | F | less than a year | SP, BP I, MD, SHB, PTSD, ICD, PSY, SI | Levetiracetam |
| 26-30 | F | 15 years | PD, AG, SAD, MD, OCD, PTSD, ICD, SD, PSY, SI, SHB | Valproate, Levetiracetam, Stiripentol, Perampanel, Pregabalin, Clobazam, Clonazepam, Oxycodone |
| 31-35 | F | 1 year | AG, SP, GAD, MD, OCD, PTSD, INS, SD, SI | Lamotrigine, Mirtazapine |

Psychiatric diagnoses are based on the structured psychiatric interview (Mini-DIPS) administered during this study. Abbreviations: CNS=central nervous system, F=Female, M=Male, PD=panic disorder, AG=agoraphobia, BP=bipolar disorder, MD=major depression, PTSD=post-traumatic stress disorder, INS=insomnia, ICD=impulse control disorder, SI=suicide ideation, SHB=self-harming behavior, SAD=social anxiety disorder, HME=hypomanic episode, PSY=psychotic episodes, GAD=generalized anxiety disorder, PDD=persistent depressive disorder, AUD=alcohol use disorder, SP=specific phobia, ED=eating disorder, SSD=somatic symptom disorder, OCD=obsessive compulsive behavior, SD=sexual dysfunction. In patients with Mini-DIPS diagnoses of panic disorder (PD), seizure semiology was distinct from panic attack symptoms
