## Supplementary Table 2 for "Linking gene expression patterns and brain morphometry to trauma and symptom severity in patients with functional seizures"

**Supplementary Table 2. Demographic, Clinical Information and Questionnaire Scores in Patients with Functional Seizures.**

| | M(SD) |
|---|---|
| **Demographics** | |
| Age (years) | 34.9 (11.2) |
| Sex (female : male) | 15 : 5 |
| Education level (years) | 11.1 (1.8) |
| **Illness severity** | |
| Illness duration (years) | 5.2 (5.3) |
| SDQ-20 | 34.7 (9.9) |
| **Adverse Life Events** | |
| TEC Overall score | 7.2 (4.81) |
| TEC Emotional neglect | 2.85 (3.12) |
| TEC Emotional abuse | 1.65 (1.63) |
| TEC Physical abuse | 1.75 (1.65) |
| TEC Threat to life | .7 (1.38) |
| TEC Sexual trauma | 2.05 (3.38) |

Abbreviations: M=mean, SD=Standard Deviation, SDQ-20=Somatic Dissociation Questionnaire, TEC=Traumatic Experiences Checklist.
