## Supplementary Table 3 for "Linking gene expression patterns and brain morphometry to trauma and symptom severity in patients with functional seizures"

**Supplementary Table 3. Full list of genes with high spatial similarity to gray matter volume correlations for different analyses.**

**a)** List of genes with high spatial similarity to gray matter volume correlation with SDQ-20 (135 genes)

| Cluster1 | | | Cluster2 | | |
|---|---|---|---|---|---|
| ABCC12 | KCNC2 | POU2F2 | ABCA3 | GNG3 | PKD2L1 |
| ANKRD13B | KCNH5 | PROKR2 | ABHD12B | GRM3 | PLCH1 |
| C1orf115 | KCNJ12 | PTPRQ | ACOT7 | GSG1L | PNMAL2 |
| C7orf51 | KCNJ6 | RAB3A | ADCY2 | HPCAL1 | PPEF1 |
| CBLN1 | KRT31 | RASGEF1C | ADCYAP1 | HTR3B | PRRT1 |
| CBLN2 | L1CAM | RASGRF2 | ARHGDIG | IGSF21 | RGS4 |
| CCDC85A | LINC00087 | RASGRP1 | ASXL3 | IGSF8 | RHBDL1 |
| CHRNB3 | LINGO4 | RGS11 | B3GNT1 | KCNB2 | RHEBL1 |
| CX3CR1 | LRFN2 | RPRM | C20orf27 | KCND2 | RIT2 |
| DBH | LRRC24 | SPRN | C6orf154 | KCNF1 | RTP1 |
| DPF1 | MCHR1 | SSPO | CA10 | KIAA1024 | RXFP1 |
| EXTL1 | MCHR2 | STX1A | CALHM1 | LILRA4 | SCG2 |
| FAM148C | MGAT4C | SYT10 | CCK | LMO3 | SEZ6L2 |
| FAM163A | MGAT5B | SYT4 | CDH18 | LMO4 | SLC1A6 |
| FIBCD1 | MPPED1 | TCERG1L | DNAH7 | LRRTM4 | SLC30A3 |
| FRMPD2 | NANOS3 | TMEM132A | DPP10 | LY6H | SVOP |
| GALNT9 | NHLH1 | TMEM132D | DYNC1I1 | MSANTD1 | SYNPR |
| GPM6A | NKX2-4 | UCMA | ESYT3 | NDST3 | TMEM121 |
| GPR26 | NRGN | WBSCR17 | FAIM2 | NETO2 | TMEM196 |
| HBE1 | PCDH10 | WFIKKN2 | FAM131A | NEUROD6 | TMEM59L |
| HDC | PCDH20 | XKR4 | FBXO44 | NGB | VAMP2 |
| HOMER1 | PNMA5 | ZIC3 | FGF20 | NOL4 | |
| HPCA | PNOC | | FREM3 | PCBP3 | |

**b)** List of genes with high spatial similarity to gray matter volume correlation with TEC Emotional neglect (121 genes)

| Cluster1 | | | Cluster2 | | |
|---|---|---|---|---|---|
| AKAP14 | GNG4 | TMEM130 | ANKRD6 | GNAO1 | PCDH20 |
| BZRAP1 | GPR88 | YPEL1 | APBA1 | GNG2 | PCDHGC3 |
| C1orf173 | GPRIN1 | ZCCHC18 | ASTN1 | GP1BB | PGM2L1 |
| C1orf187 | HPCAL4 | | ATOH7 | GPM6A | PKIB |
| C20orf27 | HTR1A | | C1orf95 | GRIA1 | PNCK |
| C2orf80 | HTR2C | | C1QL1 | GRM1 | PPP4R4 |
| CALB2 | ICAM5 | | C6orf221 | HHIPL1 | PRKCG |
| CAMKV | KCNA4 | | CAPSL | LINC00087 | PTCHD1 |
| CDH4 | KCNN3 | | CCDC37 | LRRC3B | RAB27B |
| CENPVL1 | KCTD4 | | CDH9 | LY6H | RIMBP2 |
| CHST1 | LUZP2 | | CPNE6 | MAPK1 | RTBDN |

| | | | | |
|---|---|---|---|---|
| COCH | MAGED4B | CPT1C | MAPK8IP1 | SCN3B |
| CPLX3 | NECAB2 | CTXN1 | MMD | SLC17A7 |
| CXorf57 | NNAT | CX3CR1 | MMD2 | SLC1A3 |
| DLL3 | NOL4 | CXXC11 | MSANTD1 | SLC39A12 |
| DNAH7 | ODZ3 | DMC1 | NEUROD6 | SLIT1 |
| DNAJC12 | PDYN | FAM148C | NKAIN3 | SST |
| DYDC2 | PEA15 | FAM19A1 | NKAIN4 | SSTR1 |
| FABP7 | PTPRR | FXYD6 | NPB | SYT17 |
| FAM171B | PYDC1 | GABRA3 | NPTXR | TMEM132A |
| FAM181A | RAPGEF4 | GABRA5 | NTSR1 | TUBB2A |
| GABRA2 | SCN3A | GDA | P2RY12 | ZCCHC12 |
| GABRB1 | SNCA | GLRA3 | PALM | |
| GLRA2 | THRA | GMFB | PCDH19 | |

**c)** List of genes with high spatial similarity to gray matter volume correlation with TEC Sexual trauma (108 genes)

| **Cluster1** | | **Cluster2** | | |
|---|---|---|---|---|
| ABHD12B | PLCH1 | APBA1 | DPF1 | MMP17 |
| ADCY2 | RASGRF2 | ARF3 | DPP10 | MYRIP |
| CA10 | RHEBL1 | ARHGDIG | DTNBP1 | NANOS3 |
| DYNC1I1 | RIT2 | ASXL3 | EFNB3 | NAV3 |
| FAM133A | RTP1 | B3GNT1 | FAM131A | NETO1 |
| FGF20 | RXFP1 | C20orf27 | FAM148C | NEUROD6 |
| FJX1 | SH2D5 | C2orf80 | FRMPD2 | NKX2-4 |
| FREM3 | SLC30A3 | C6orf154 | GAP43 | NOL4 |
| GNG3 | SLC9A6 | C8orf46 | GDAP1L1 | PCDH20 |
| GRIA4 | TMEM196 | CA11 | GNG2 | PITPNM3 |
| GRM3 | TMEM59L | CAMKV | GRIK1 | PPP4R4 |
| HTR1E | TRPC5 | CBLN1 | GSG1L | RAB15 |
| HTR3B | | CD86 | HBQ1 | RASGEF1C |
| ITPKA | | CDH10 | HPCAL1 | RILPL1 |
| KCND2 | | CDH4 | HSD11B1L | SCG2 |
| KCNN2 | | CDH9 | KCNB2 | SNCA |
| KCNV1 | | CGREF1 | KCNG3 | SPRN |
| KIF17 | | CHST1 | KCNMB4 | SVOP |
| LRRTM4 | | CPNE5 | KIAA1024 | SYNPR |
| NDST3 | | CTNND2 | LASS1 | SYT5 |
| NETO2 | | DACH2 | LILRA4 | TFPT |
| NPY5R | | DDN | LINC00087 | VAMP2 |
| PCDH8 | | DNAH7 | LRRC16B | YJEFN3 |
| PKD2L1 | | DOC2A | LY6H | YWHAH |

**d)** List of genes overlapping between SDQ-20 Cluster 2 (genes listed in Supplementary Table 4a) and TEC Sexual Trauma Cluster 1 (genes listed in Supplementary Table 4c) (22 genes)

| Overlapping | |
|---|---|
| ABHD12B | NDST3 |
| ADCY2 | NETO2 |
| CA10 | PKD2L1 |
| DYNC1I1 | PLCH1 |
| FGF20 | RHEBL1 |
| FREM3 | RIT2 |
| GNG3 | RTP1 |
| GRM3 | RXFP1 |
| HTR3B | SLC30A3 |
| KCND2 | TMEM196 |
| LRRTM4 | TMEM59L |

**e)** List of genes overlapping between SDQ-20 Cluster 2 (genes listed in Supplementary Table 4a) and TEC Emotional neglect Cluster 2 (genes listed in Supplementary Table 4b) (3 genes)

| Overlapping |
|---|
| C20orf27 |
| DNAH7 |
| NOL4 |
