## Supplementary Table 4 for "Linking gene expression patterns and brain morphometry to trauma and symptom severity in patients with functional seizures"

**Supplementary Table 4. Full list of PANTHER Pathways associated with the genes with high spatial similarity to GM volume correlations for different analyses.**

**a)** Full list of PANTHER Pathways associated with the genes with high spatial similarity to gray matter volume correlation with SDQ-20 Cluster 2, surviving to FDR correction (genes listed in Supplementary Table 4a)

| **PANTHER Pathways** | **Fold Enrichment** | **p (FDR)** |
|---|---|---|
| 5HT3 type receptor mediated signaling | 32.36 | 0.02 |
| Beta3 adrenergic receptor signaling | 30.74 | 0.03 |
| 5HT4 type receptor mediated signaling | 26.35 | 0.02 |
| Opioid prodynorphin | 25.62 | 0.02 |
| Opioid proopiomelanocortin | 24.92 | 0.01 |
| Opioid proenkephalin | 24.92 | 0.01 |
| Histamine H2 receptor mediated signaling | 24.59 | 0.03 |
| Beta2 adrenergic receptor signaling | 19.62 | 0.02 |
| Beta1 adrenergic receptor signaling | 19.62 | 0.01 |
| 5HT1 type receptor mediated signaling | 19.21 | 0.01 |
| Metabotropic glutamate receptor group II | 18.82 | 0.01 |
| Ionotropic glutamate receptor | 18.44 | 0.01 |
| Dopamine receptor mediated signaling | 15.63 | 0.02 |
| Oxytocin receptor mediated signaling | 15.37 | 0.02 |
| Thyrotropin-releasing hormone receptor signaling | 14.87 | 0.02 |
| 5HT2 type receptor mediated signaling | 13.36 | 0.02 |
| Metabotropic glutamate receptor group III | 13.17 | 0.02 |
| Heterotrimeric G-protein signaling-Gi alpha and Gs alpha mediated | 7.41 | 0.02 |

**b)** Full list of PANTHER Pathways associated with the genes with high spatial similarity to gray matter volume correlation with TEC Emotional neglect Cluster 2, surviving to FDR correction (genes listed in Supplementary Table 4b)

| **PANTHER pathways** | **Fold Enrichment** | **p (FDR)** |
|---|---|---|
| Metabotropic glutamate receptor group III | 21.63 | 0.00 |
| Heterotrimeric G-protein signaling-Gq alpha and Go alpha mediated | 12.21 | 0.01 |
| Wnt signaling | 6.69 | 0.01 |

**c)** Full list of PANTHER Pathways associated with the genes with high spatial similarity to gray matter volume correlation with TEC Sexual trauma Cluster 1, surviving to FDR correction (genes listed in Supplementary Table 4c)

| **PANTHER pathways** | **Fold Enrichment** | **p (FDR)** |
|---|---|---|
| 5HT1 type receptor mediated signaling | 35.76 | 0.02 |
| 5HT4 type receptor mediated signaling | 32.69 | 0.05 |
| GABA-B receptor II signaling | 31.78 | 0.05 |
| Opioid prodynorphin | 31.78 | 0.04 |
| Opioid proopiomelanocortin | 30.92 | 0.04 |
| Opioid proenkephalin | 30.92 | 0.04 |
| Histamine H1 receptor mediated signaling | 26.00 | 0.04 |
| Beta2 adrenergic receptor signaling | 24.34 | 0.05 |
| Beta1 adrenergic receptor signaling | 24.34 | 0.04 |
| Metabotropic glutamate receptor group II | 23.35 | 0.04 |
| Ionotropic glutamate receptor | 22.88 | 0.04 |
| Dopamine receptor mediated signaling | 19.39 | 0.05 |
| Oxytocin receptor mediated signaling | 19.07 | 0.05 |
| Thyrotropin-releasing hormone receptor signaling | 18.45 | 0.05 |
| Heterotrimeric G-protein signaling-Gi alpha and Gs alpha mediated | 13.79 | 0.02 |

**d)** Full list of PANTHER Pathways associated with the genes overlapping between SDQ-20 Cluster 2 and TEC Sexual Trauma Cluster 1, surviving to FDR correction (genes listed in Supplementary Table 4d)

| **PANTHER Pathways** | **Fold Enrichment** | **p (FDR)** |
|---|---|---|
| Beta3 adrenergic receptor signaling | 62.41 | 0.04 |
| Enkephalin release | 53.49 | 0.04 |
| 5HT4 type receptor mediated signaling | 53.49 | 0.03 |
| GABA-B receptor II signaling | 52.01 | 0.02 |
| Opioid prodynorphin | 52.01 | 0.02 |
| Opioid proopiomelanocortin | 50.60 | 0.02 |
| Opioid proenkephalin | 50.60 | 0.01 |
| Histamine H1 receptor mediated signaling | 42.55 | 0.02 |
| Beta2 adrenergic receptor signaling | 39.84 | 0.02 |
| Beta1 adrenergic receptor signaling | 39.84 | 0.02 |
| 5HT1 type receptor mediated signaling | 39.01 | 0.02 |
| Metabotropic glutamate receptor group II | 38.21 | 0.02 |
| Dopamine receptor mediated signaling | 31.73 | 0.02 |
| Oxytocin receptor mediated signaling | 31.20 | 0.02 |
| Thyrotropin-releasing hormone receptor signaling | 30.20 | 0.02 |
| 5HT2 type receptor mediated signaling | 27.13 | 0.02 |

| | | |
|---|---|---|
| Heterotrimeric G-protein signaling -Gi alpha and Gs alpha mediated | 16.92 | 0.02 |
| Inflammation mediated by chemokine and cytokine signaling | 11.01 | 0.02 |
