## Supplementary Table 5 for "Linking gene expression patterns and brain morphometry to trauma and symptom severity in patients with functional seizures"

**Supplementary Table 5. Information on clusters remaining significant after whole-brain, cluster-wise correction for multiple comparisons with a cluster-forming threshold p<.05**

| Contrast of interest | Corresponding figure | Region of cluster peak | Max T-value | Peak p-value (uncorrected) | Cluster size ($mm^2$) |
|---|---|---|---|---|---|
| SDQ-20 | Figure 2A | L Insula/Precentral | -6.12 | <.001 | 8160 |
| | | L Rostral Anterior Cingulate | -4.47 | <.001 | 7480 |
| | | R Insula | -5.34 | <.001 | 5768 |
| | | L Lateral Orbitofrontal | -4.29 | <.001 | 3408 |
| | | Brainstem | -6.00 | <.001 | 3288 |
| | | R Isthmus Cingulate | -4.22 | <.001 | 3024 |
| | | R Precentral | -6.17 | <.001 | 1600 |
| Illness duration | Figure 2B | L Caudal Anterior Cingulate/Superior Frontal | -4.51 | <.001 | 4608 |
| | | R Insula/Pars Opercularis | -5.97 | <.001 | 4392 |
| Emotional neglect | Figure 3A | R Fusiform | 5.42 | <.001 | 4648 |
| | | L Insula | -6.30 | <.001 | 3624 |
| | | R Amygdala/Inferior Temporal | -4.57 | <.001 | 3104 |
| | | R Insula/Putamen | -4.75 | <.001 | 3096 |

Abbreviations: FS=Functional seizures, L=Left, R=Right, SDQ-20=Somatoform Dissociation Questionnaire.
