## Supplementary Figure 1 for "Linking gene expression patterns and brain morphometry to trauma and symptom severity in patients with functional seizures"

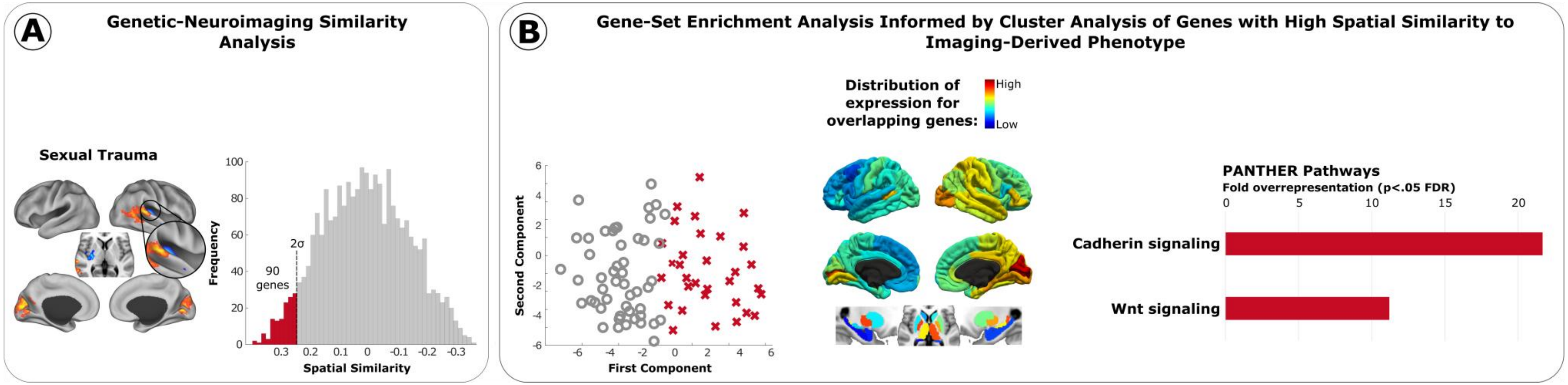


**Supplementary Figure 1. Gene-Expression Analysis: Relationship between gray matter volume – sexual trauma relationships and enriched gene-sets based on spatial similarity.** Genetic-neuroimaging similarity analyses between gene expression and gray matter, as well as gene-set enrichment analyses were computed for the map derived from sexual trauma (positive tail). **Panel A** displays the spatial similarity analysis: The Imaging-derived phenotype (t-statistic map) for the association between gray matter volume and sexual trauma was correlated with spatial expression maps from the AHBA. Genes with a spatial similarity >2 SD from the mean in the upper tail of the distribution were extracted. **Panel B** shows the Gene-set enrichment: Cluster analysis revealed two clusters for the spatial similarity distribution (left side of panel B). The distribution of gene expression for the genes from the colored cluster was mapped to enable a cross-reference to the original imaging-derived phenotype (middle of panel B). The list of genes for the cluster was entered into gene-set enrichment analyses for PANTHER pathways (right side of panel B).
